# Examining the inter-relationships between social isolation and loneliness and their correlates among older British adults before and during the COVID-19 lockdown: evidence from four British longitudinal studies

**DOI:** 10.1101/2023.06.27.23291947

**Authors:** Rosie Mansfield, Giorgio Di Gessa, Kishan Patel, Eoin McElroy, Jaques Wels, Morag Henderson, Jane Maddock, Jean Stafford, Andrew Steptoe, Marcus Richards, Praveetha Patalay

## Abstract

**Background and Objectives:** Unprecedented social restrictions during the COVID-19 pandemic have provided a new lens for considering the inter-relationship between social isolation and loneliness in later life. We present these inter-relationships before and during the COVID-19 restrictions and investigate to what extent demographic, socio-economic, and health factors associated with such experiences differed during the pandemic.

**Research Design and Method:** We used data from four British longitudinal population-based studies (1946 MRC NSHD, 1958 NCDS, 1970 BCS, and ELSA). Rates, co-occurrences, and correlates of social isolation and loneliness are presented prior to and during the early stage of the COVID-19 pandemic and the inter-relationships between these experiences are elucidated in both periods.

**Results:** Across the four studies, pre-pandemic proportions reporting social isolation ranged from 15 to 54%, with higher rates in older ages (e.g., 32% of 70-79 and 54% of those over 80). During the pandemic, the percentage of older people reporting both social isolation and loneliness and isolation only slightly increased. The inter-relationship between social isolation and loneliness did not change. Associations between socio-demographic and health characteristics and social isolation and loneliness also remained consistent, with greater burden among those with greater economic precarity (females, non-homeowners, unemployed, illness and greater financial stress).

**Discussion and Implications:** There were already large inequalities in experiences of social isolation and loneliness and the pandemic had a small impact on worsening these inequalities. The concepts of loneliness and social isolation are not transferable and clarity is needed in how they are conceptualised, operationalised, and interpreted.

## Background and Objectives

Prior to the COVID-19 pandemic, loneliness was identified as a significant public health concern (Jeste et al., 2019): for example, in the UK, a Minister for Loneliness was appointed in 2018 and the ’Campaign to End Loneliness’ was launched. Despite increased policy interest, there remains a need to better understand the conceptualisation and measurement of social isolation and loneliness as they are often inconsistently applied and interchangeably referred to across research, policy, and practice (Wigfield, 2020). With the implementation of social distancing and quarantine measures due to the COVID-19 pandemic and several national lockdowns across Britain, social isolation – and its relationship with loneliness – were brought into even sharper focus. By carefully considering the operationalisation of these complex concepts, this study has the dual aim to 1) explore the conceptual and empirical inter-relationships between social isolation and loneliness, and 2) identify overlapping and independent correlates of social isolation and loneliness prior to and during the COVID-19 pandemic. Using data from four British longitudinal studies, we investigate associations between social isolation and loneliness under normal circumstances and during COVID-19 restrictions and examine whether the factors associated with such experiences differ due to the pandemic.

As an objective condition, social isolation can be quantified by a person’s network size, composition, and frequency of contact (Holt-Lunstad et al., 2015; Huisman & van Tilburg, 2021). On the other hand, the way in which an individual perceives and experiences their social circumstances includes qualitative assessments of the value, function, and meaning ascribed to relationships. Loneliness arises as a negative feeling associated with the perception of an inadequate quantity and or quality of social relationships (Zavaleta et al., 2017). It can therefore be experienced in the absence of isolation and vice versa, i.e., those who are socially isolated may not experience loneliness (Dykstra, 2006; Perlman & Letitia Anne Peplau, 1984; J. E. M. Power et al., 2019)

Data from the European Social Survey collected before the pandemic indicated that 8.6 percent of the adult population had frequent feelings of loneliness whereas 20.8 percent were socially isolated (d’Hombres et al., 2021). It is clear that one experience can exist without the other, with only a moderate association observed between social isolation and loneliness (Hughes et al., 2004). Both constructs have also been found to independently predict poorer health, wellbeing, cognitive capability, and mortality in older age through different mechanisms (Coyle & Dugan, 2012; Golden et al., 2009; Steptoe, Shankar, et al., 2013), providing empirical evidence for a conceptual distinction between these two constructs. However, the investigation of objective indicators of social isolation is often neglected, with few studies examining the interaction between isolation and loneliness (Holt-Lunstad & Steptoe, 2022).

The burden of these experiences is also not equally shared and, although overlapping, the socio-demographic factors associated with social isolation and loneliness are varied. Common risk factors include low economic position and poor health; however, older age is associated with increased social isolation but not loneliness (d’Hombres et al., 2021). Despite being more likely to live alone in later life, recent findings indicate that women are less lonely than men (Barreto et al., 2021; Esteve et al., 2020). Investigations of the interaction between age and other socio-demographic factors provide a more nuanced picture. For example, low education level, deprivation, and female gender were only associated with loneliness in adolescence and early adulthood in a large Danish population-based study (Lasgaard et al., 2016). In the UK, lower levels of loneliness were associated with the number of social interactions in early adult life and relationship quality in midlife (Victor & Yang, 2012).

Although several cross-sectional studies have indicated high levels of social isolation and loneliness during the COVID-19 restrictions, it is difficult to infer causality in the absence of pre-pandemic scores (Killgore et al., 2020). The first large-scale, population-based study investigating the psychological impact of the pandemic was based on data collected in the first Understanding Society COVID-19 survey (Li & Wang, 2020). Over a third of the sample reported feeling lonely sometimes or often during the pandemic. Young people, women, and those with COVID-19 symptoms were more likely to report loneliness and mental health difficulties, while those in employment and with a cohabiting partner were found to report less loneliness. Low income, not being married or cohabiting, smaller household size (adults only), higher depressive symptoms, living in an urban area, and lower number of close friends and social support were also associated with loneliness (Bu et al., 2020; Groarke et al., 2020).

Limited to the unique experience of lockdown, few of these studies tell us much about the stability of demographic, socio-economic, and health characteristics associated with social isolation and loneliness before and during the pandemic and the strength of these associations. Were the people at risk of social isolation and loneliness prior to COVID-19 more likely to have these experiences during lockdown? Or were new groups disproportionately affected by the drastic changes to their lifestyle? To answer these questions in relation to loneliness, Bu et al., (2020) conducted a cross-cohort study using data from the UK Household Longitudinal Study (Understanding Society) (2017-2019) and the COVID-19 Social Study. Different groups of individuals were identified, including those whose risk of loneliness remained the same during the pandemic (e.g., women, urban residents, and those living alone), those who experienced heightened risk (e.g., those with low income and young people), and those who emerged as high-risk groups during the pandemic (e.g., students). Due to the use of different cohorts, this study is unable to directly compare the experiences of individuals over time and therefore cannot make conclusions about changes due to the pandemic. The use of multiple successive birth cohorts in the current study, alongside a multi-generational ageing cohort, provides a further opportunity to examine cross-generational differences in experiences during the pandemic and tease apart age or cohort effects from period effects related to the pandemic.

This article aims to overcome some of the existing methodological limitations in the field and has two main aims. First, we describe levels of social isolation and loneliness prior to and during the COVID-19 restrictions, testing associations between social isolation and loneliness under normal circumstances and during the early stage of COVID-19 restrictions. By following the same individuals before and during the pandemic, we also provide better evidence for the differential impacts of restrictions on social isolation, loneliness, and their intersection. Second, we compare the correlates of social isolation and loneliness, and the relationship between them, pre- and during the COVID-19 restrictions to answer the following research questions:

### Research Questions

1. What were the levels of social isolation and loneliness, and what proportion of the sample was classified into different groups e.g., isolated, and/or lonely prior to and during the COVID-19 restrictions?
2. What were the inter-relationships between social isolation and loneliness indicators prior to and during the COVID-19 restrictions?
3. To what extent were demographic, socio-economic factors, and physical and mental health associated with social isolation and loneliness prior to and during the COVID-19 restrictions?

## Research Design and Methods

### Data sources

Data collected at two time points prior to and during the COVID-19 pandemic were utilised from four UK population-based studies. Three are longitudinal birth cohort studies with samples born within a single week across England, Scotland, and Wales: the 1946 MRC National Survey of Health and Development (NSHD) (Kuh et al., 2016; Wadsworth et al., 2006), the 1958 National Child Development Study (NCDS) (C. Power & Elliott, 2006), and the 1970 British Cohort Study (BCS) (Elliott & Shepherd, 2006; Sullivan et al., 2022)). In contrast, the English Longitudinal Study of Ageing (ELSA) is a panel study following individuals aged ≥50 years biennially since 2002 (Steptoe, Breeze, et al., 2013).

Table 1 provides details of the studies including participants’ age and timing of data collection across surveys. It also provides details of the survey designs, sampling frames, response rates, and analytic sample sizes.

**Table 1.**
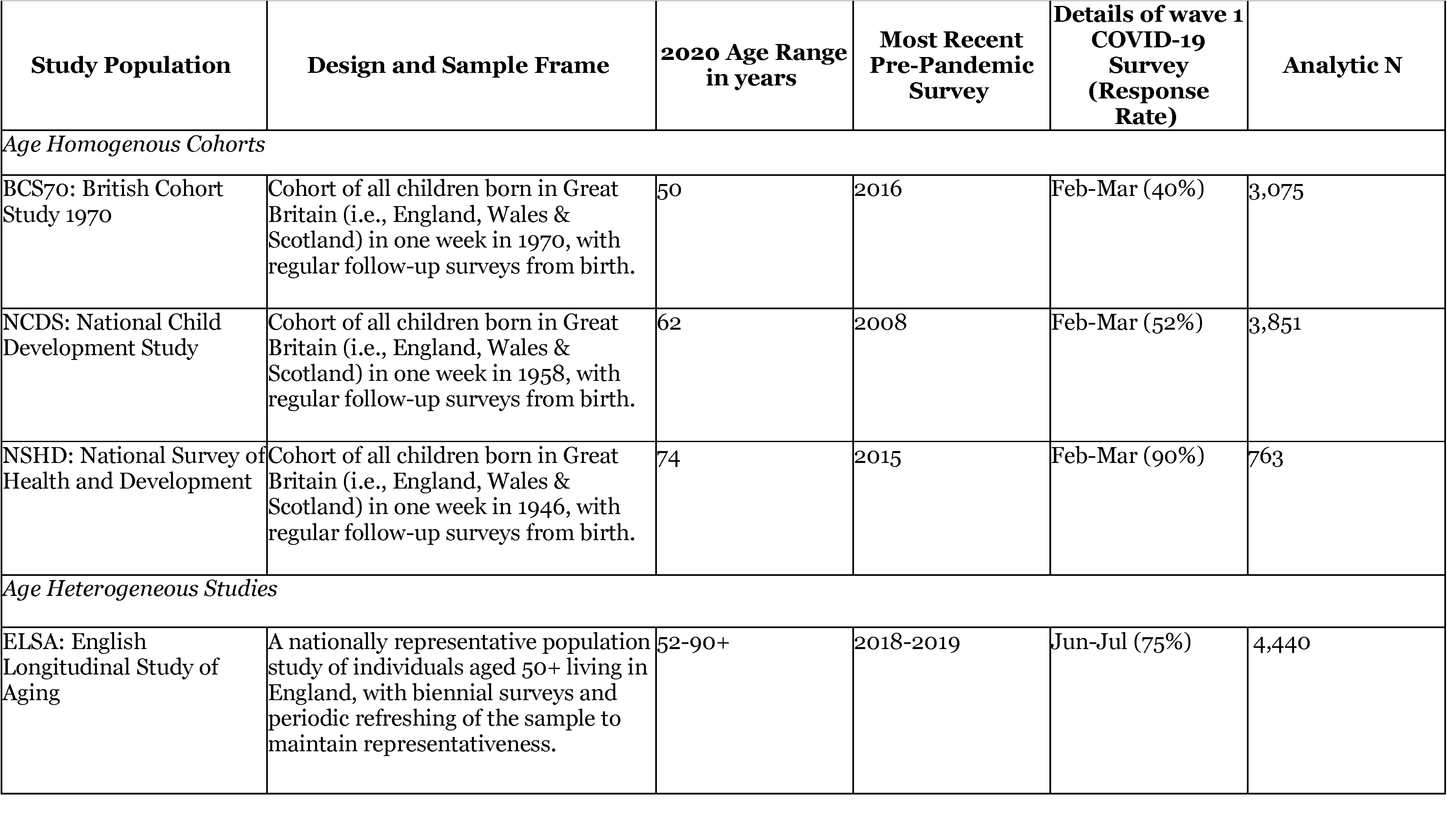
Details of the cohort studies including participants’ age and timing of data collection, survey designs, sampling frames, response rates, and analytic sample sizes

### Participants

The analytic sample for each cohort included those who were alive, living in Great Britain, who took part in the study at two time points prior to and one wave during the early stages of the pandemic, and who completed the pre-pandemic and wave 1 COVID-19 surveys including outcomes of interest (social isolation and loneliness). Across all cohorts, we further restricted the sample to those who directly participated in the surveys (i.e. we excluded proxy respondents). Age bands that mapped onto the age homogenous cohorts during the COVID-19 pandemic (e.g., 50-59, 60-69, 70-79, and 80+) were generated for ELSA participants to enable analysis to differentiate between age or cohort and period effects related to the pandemic. Participants’ demographic, socio-economic, and health characteristics are summarised in <u>supplementary file S1</u>.

### Sampling Design Weights and Accounting for Non-Response Bias

To account for sampling design and non-response biases, weights were applied to studies to improve representativeness of their target populations i.e., the general population of mid to older age adults in Great Britain/England. For NSHD, design weights were included in the generation of non-response weights for the wave 1 COVID-19 survey. Only survey non-response weights for the wave 1 COVID-19 timepoint were included for NCDS, BCS, and ELSA. Given that certain groups of individuals are more likely to discontinue participation in longitudinal surveys (e.g., males and those disadvantaged and less healthy), accounting for non-response in analyses ensures that data from these participants are given more weight, resulting in a more representative sample.

### Measures

This section provides details of the measurement of social isolation and loneliness prior to and during the COVID-19 pandemic, and an overview of demographic, socio-economic, and health characteristics being investigated in the current study. Full details of the original items, harmonisation, and re-coding are included in <u>supplementary file S2</u>.

#### Social isolation

Self-reported indicators of social isolation were identified in the four studies prior to and during the COVID-19 restrictions. Relevant items were organised by their relational context (e.g., household, community) and by the domain of social isolation assessed (e.g., network size, frequency of contact). For example, items were generated for isolation within the household (i.e., living alone), family network (i.e., partnership and children), frequency of contact with friends and relatives, education and employment status, frequency of contact with people in the community (e.g., frequency of attending community groups/organisations), and volunteering. To compare study members who were isolated across contexts prior to and during the COVID-19 restrictions, items were reduced to those which could be harmonised across time points. An overall social isolation score was generated with a total index score (maximum value of six) indicating the number of contexts an individual is isolated across. This variable was also recoded as a binary variable, for estimating proportions and visualising overlaps with loneliness, where a score greater than three (indicating isolation in at least three contexts) was used as a binary indicator of social isolation.

#### Loneliness

Prior to the COVID-19 pandemic, not all cohorts asked participants a full measure of loneliness such as the UCLA Loneliness Scale. However, items included ‘I feel left out of things’ and related to the extent to which cohort members had been feeling close to others e.g., ‘I’ve been feeling close to other people’. To make variables comparable across cohorts, items were recoded as binary, indicating those that were lonely and not lonely. Across all four cohorts, the UCLA Loneliness Scale (3-item) (Hughes et al., 2004) was included in the COVID-19 survey along with an overall item ‘How often do you feel lonely?’. The short version of the Revised UCLA Loneliness Scale (R-UCLA; (Russell et al., 1980) consists of 3 items relating to lacking companionship, feeling left out, and feeling isolated from others, with simplified response options (‘*hardly ever*’ = 1, ‘*some of the time*’ = 2, or ‘*often*’ = 3). For cohorts that did not include the UCLA Loneliness Scale prior to the COVID-19 pandemic, the best-matched item was selected during COVID-19 to generate the loneliness indicator. For example, as can be seen in <u>supplementary file S2</u>, in NCDS there was only 1 item relating to feeling left out collected prior to the COVID-19 pandemic; this was therefore matched with the UCLA item during COVID-19 relating to feeling left out to generate the most comparable loneliness indicators. Designed for large-scale social surveys, the three-item UCLA Loneliness Scale provides a reliable indicator of loneliness. assessment of loneliness. Only a modest relationship was found between this measure of loneliness and objective social isolation (Hughes et al., 2004) supporting the conceptual distinction in the current study.

#### Demographic, socio-economic and health characteristics

Demographic variables included sex as well as age and ethnicity (in ELSA only). Cohort members’ highest level of educational achievement (degree vs. no degree) was also included. Socio-economic indicators were self-reported financial difficulties, home ownership, and occupational social class. A binary variable was generated to indicate those with ‘*poor/fair*’ health (vs good, very good, or excellent health). In addition, whether cohort members report a limiting longstanding illness or health problem was included as a binary variable. Continuous measures of psychological distress and life satisfaction were also included as indicators of mental health and wellbeing (see <u>Supplementary File S2</u> for details).

### Analysis strategy

To answer the first research question and investigate the percentage of participants experiencing social isolation and loneliness prior to and during the first COVID-19 lockdown, binary variables were generated to identify those in each group. Using these indicator variables, the percentage of the sample reporting both, one, or neither are summarised prior to and during the COVID-19 restrictions. Analyses were stratified using age bands that mapped onto the other cohorts during the wave 1 COVID-19 timepoint for ELSA (50-59, 60-69, 70-79, 80+). We also present Venn diagrams depicting the proportion of the cohort experiencing social isolation and loneliness prior to and during the pandemic and the extent of overlap between these experiences.

Our second research question was addressed by examining the associations between individual indicators of social isolation and loneliness prior to and during the first COVID-19 lockdown using tetrachoric correlations. Within each cohort, we estimated tetrachoric correlation matrices using data collected prior to the pandemic, and again during the COVID-19 restrictions. We then visualised these matrices as networks, using the using ‘qgraph’ package (Epskamp et al., 2018) in R. Our nodes were all the binary indicators of social isolation and loneliness that were available in each cohort, and the edges in the networks represented the strength of the tetrachoric correlations. We chose to present bivariate relationships in the networks rather than partial correlation coefficients to avoid introducing biased or spurious connections due to inappropriate statistical control (Wysocki et al., 2022).

The extent to which demographic, socio-economic and health characteristics were associated with social isolation and loneliness prior to and during the COVID-19 pandemic was examined using two modified Poisson regression models (Zou, 2004; Zou & Donner, 2013) for each outcome of interest and within each cohort study i.e., with social isolation and loneliness as dependent variables. For ELSA, models were stratified by age bands to ensure any age effects identified in the other cohort studies were not masked by ELSA’s age heterogenous sample.

## Results

### RQ1: Descriptives and overlap of social isolation and loneliness prior to and during the COVID-19 restrictions

Table 2 provides the count and weighted percentages of the sample reporting different indicators of social isolation and loneliness prior to and during the COVID-19 restrictions. Overall rates of social isolation and loneliness are also presented for each cohort and, for ELSA, presented by age band. In addition, Table 3 summarises the count and weighted percentages of the sample reporting both social isolation and loneliness, social isolation only, loneliness only, or neither for the periods prior to and during the COVID-19 restrictions. By matching the appropriate ELSA age band with each of the other birth cohorts, Figure 1 also offers the opportunity to differentiate between age or cohort and period effects related to the pandemic.

**Figure 1.**
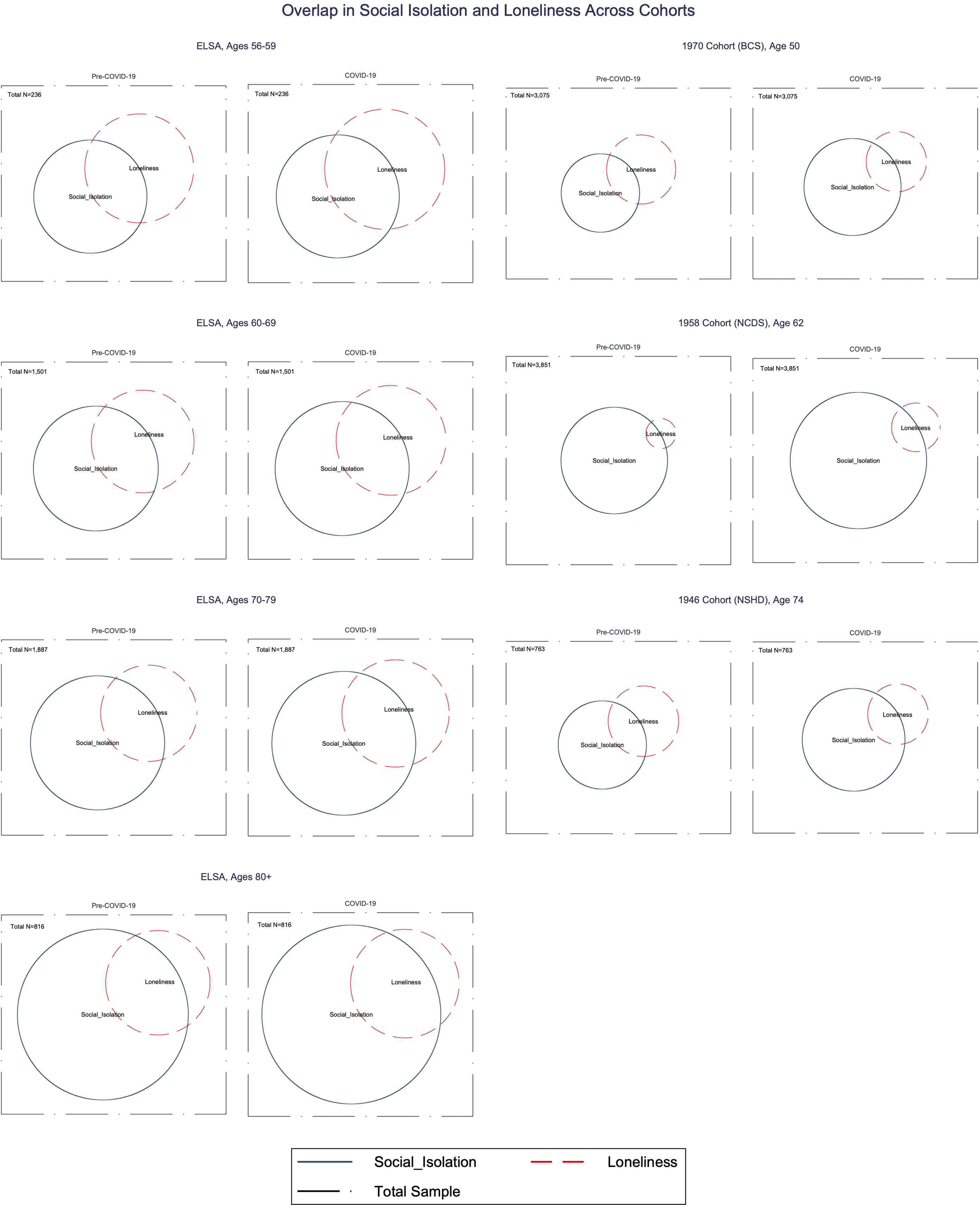
Venn diagrams

**Table 2.**
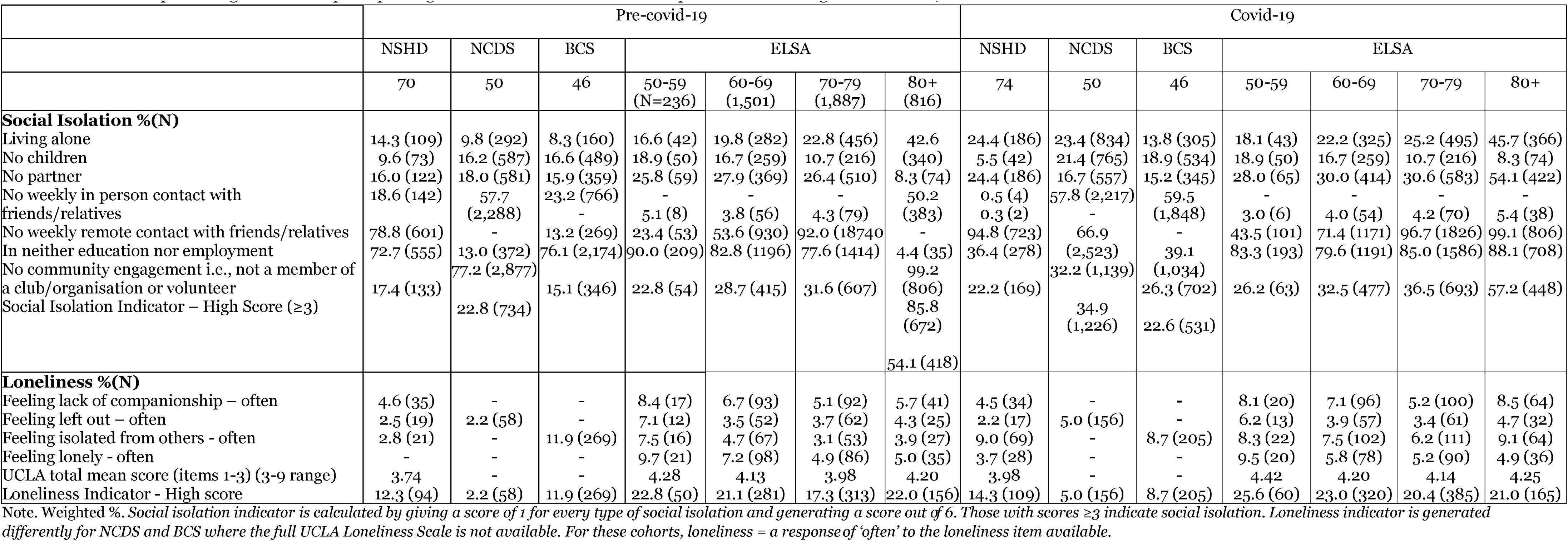
Count and percentage of the sample reporting social isolation and loneliness prior to and during the COVID-19 restrictions for each cohort

**Table 3.**
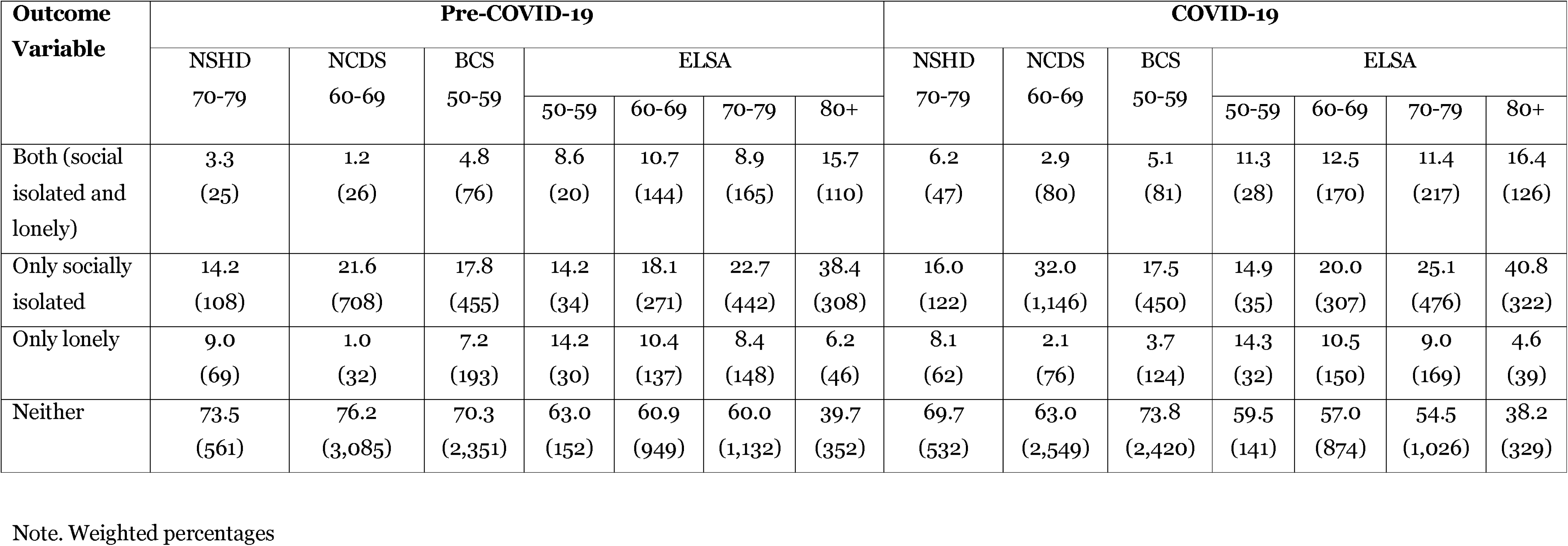
Count and % of the sample reporting possible combinations of social isolation and loneliness prior to and during the COVID-19 restrictions for each cohort and for the samples combined

Table 3 shows that the percentages of respondents reporting social isolation and loneliness is generally higher in older ages in ELSA. We observe that 8.6, 10.7, 8.9 and 15.7 percent of those aged respectively 50-59, 60-69, 70-79 and 80+ reported being both lonely and isolated in the pre-Covid-19 ELSA sample. Figures slightly increased in the Covid-19 wave with, respectively, 11.3, 12.5, 11.5 and 16.4 percent of the sample. Looking at BCS, NCDS and NSHD, percentages were 4.8, 1.2 and 3.3 in the pre-Covid-19 sweeps against 5.1, 2.9 and 6.2 during the pandemic.

The same trend is observed when looking at being socially isolated in ELSA: 14.2, 18.1, 22.,7 and 38.4 percent of those aged 50-59, 60-69, 70-7,9 and 80+ respectively reported being socially isolated in the pre-pandemic wave against 14.9, 20, 25.1 and 40.8 during the pandemic. Differences between pre-pandemic and pandemic times are of the same nature in BCS (17.8 and 17.5 percent) and NSHD (14.2 and 16 percent) but are slightly higher in NCDS (21.6 and 32 percent)

The percentages of those reporting being lonely but not socially isolated and neither socially isolated nor lonely decreased by age and did not change noticeably during the pandemic.

Finally, the percentages of those reporting being neither lonely nor socially isolated tend to be lower among older age groups. In ELSA, we can observe that 63 percent of those aged 50-59 reported neither social isolation nor loneliness against 59.5 percent during the pandemic. By contrast, figures for those aged 80+ were 39.7 percent before the pandemic and 38.2 percent during the pandemic. Percentages from BCS, NCDS and NSHD are higher but show a similar pattern with respectively 70.3, 76.3 and 73.5 percent in pre-pandemic times and 73.8, 63 and 69.7 percent during the pandemic restrictions.

The Venn diagrams in Figure 1 illustrate the overlap between isolation and loneliness for each age-group and study as well as before and during pandemic restrictions. The left side of the figures shows the proportions for each age-band within ELSA while their corresponding age-bands in BCS, NCDS and NSHD are shown on the right side. Three main observations flow from these figures. First, the size of the circles representing social isolation and loneliness combined as well as their intersections tend to be bigger within ELSA than within the birth cohorts due to higher percentage of respondents reporting neither loneliness nor social isolation within the birth cohorts. Second, the size of the intersections (i.e., those reporting being lonely and isolated) has not noticeably changed during the pandemic restrictions. Finally, it can be observed that the share of the population aged 80+ reporting being only lonely (and not isolated) is small compared with other age-bands. For those aged 80 and over, social isolation seems to be associated with loneliness more than in the other age-groups.

### RQ2: inter-relationships between social isolation and loneliness indicators prior to and during the COVID-19 restrictions

Tetrachoric correlations between all indicators of social isolation and loneliness in the NCDS and BCS are presented as networks in Figure 2, and networks by age-bands in ELSA are presented in the <u>supplementary file S3</u>. Networks could not be estimated in NSHD due to a non-positive definite correlation matrix, likely due to a tetrachoric correlation of 1 between living and partner status (i.e., all cohort members who lived alone also had no partner – likely a result of the advanced age of the cohort). As such, NSHD was excluded from this portion of the analyses. Within the NCDS and BCS cohorts, and also within ELSA age-bands, the networks were broadly similar pre- and during COVID, particularly for the strongest edges. In NCDS and BCS, loneliness was directly correlated with all measures of social isolation prior-to and during the pandemic. Prior to the pandemic, loneliness was most strongly associated with being out of work/education (NEET), living alone and having less than weekly contact with friends in the NCDS. During the pandemic, having no partner was most strongly associated with loneliness in this cohort, followed by living alone and NEET. For the BCS, the strongest pre-pandemic correlates of loneliness were living alone, NEET, and no community engagement. Living alone, having no partner, and NEET were the strongest correlates of loneliness in BCS during the pandemic.

**Figure 2.**
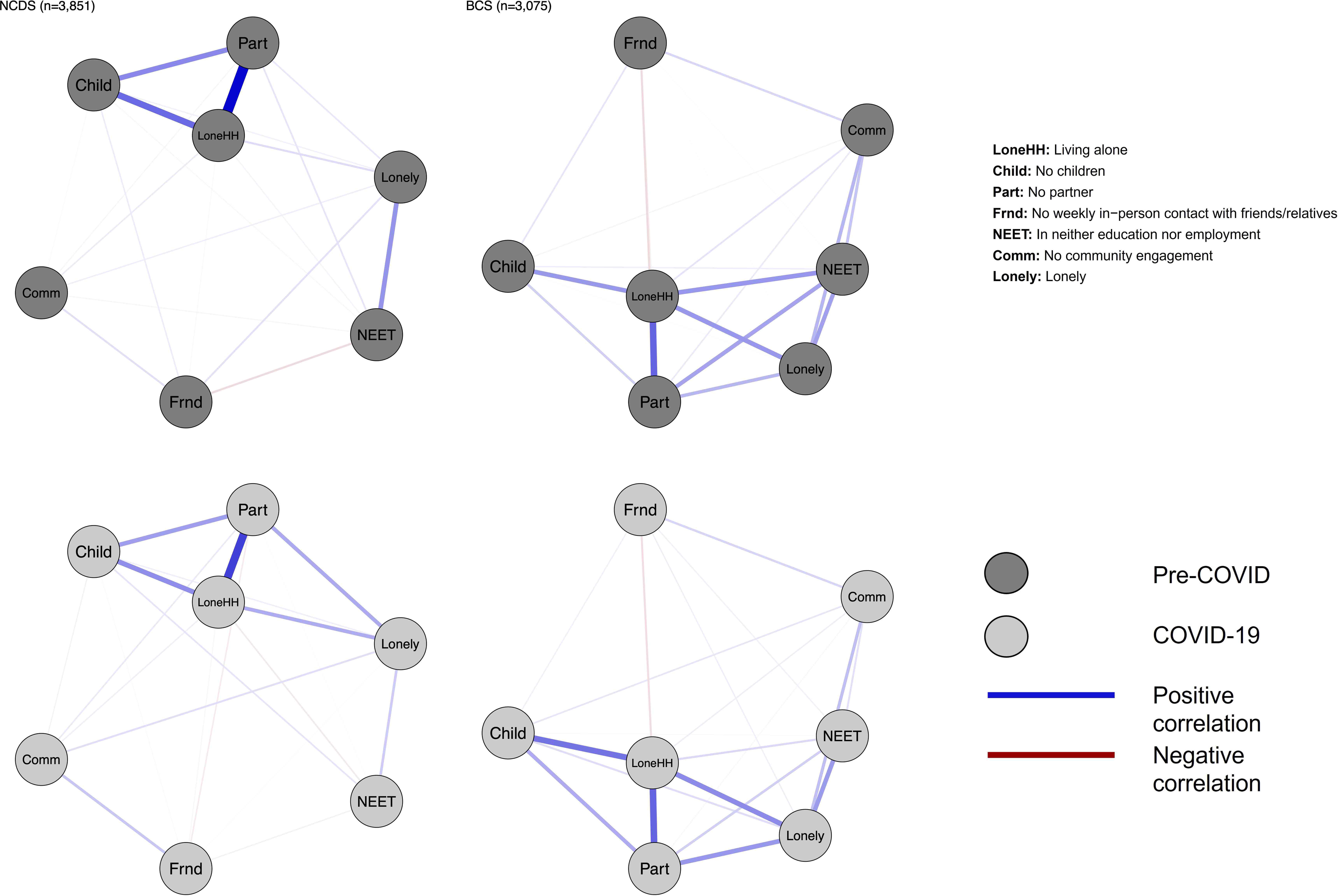
Tetrachoric correlation networks between indicators of social isolation and loneliness in NCDS and BCS

In ELSA, the four indicators of self-reported loneliness/social isolation formed a strongly connected cluster of nodes. However, these clusters had many connections with the objective indicators of social isolation. Both prior to and during the pandemic, the strongest bridges between objective and subjective indicators of social isolation were between the ‘lives alone’, ‘has no partner’, ‘lacks companionship’, and ‘feels lonely’ nodes. The tetrachoric correlation matrices used to create these networks are available in the <u>supplementary file S4.</u>

### RQ3: Predictors of social isolation and loneliness prior to and during the COVID-19 restrictions

Table 4 presents the results from the modified Poisson regression models for social isolation and loneliness prior to and during the COVID-19 pandemic. Demographic, socio-economic and health variables were added into the model in blocks.

**Table 4.**
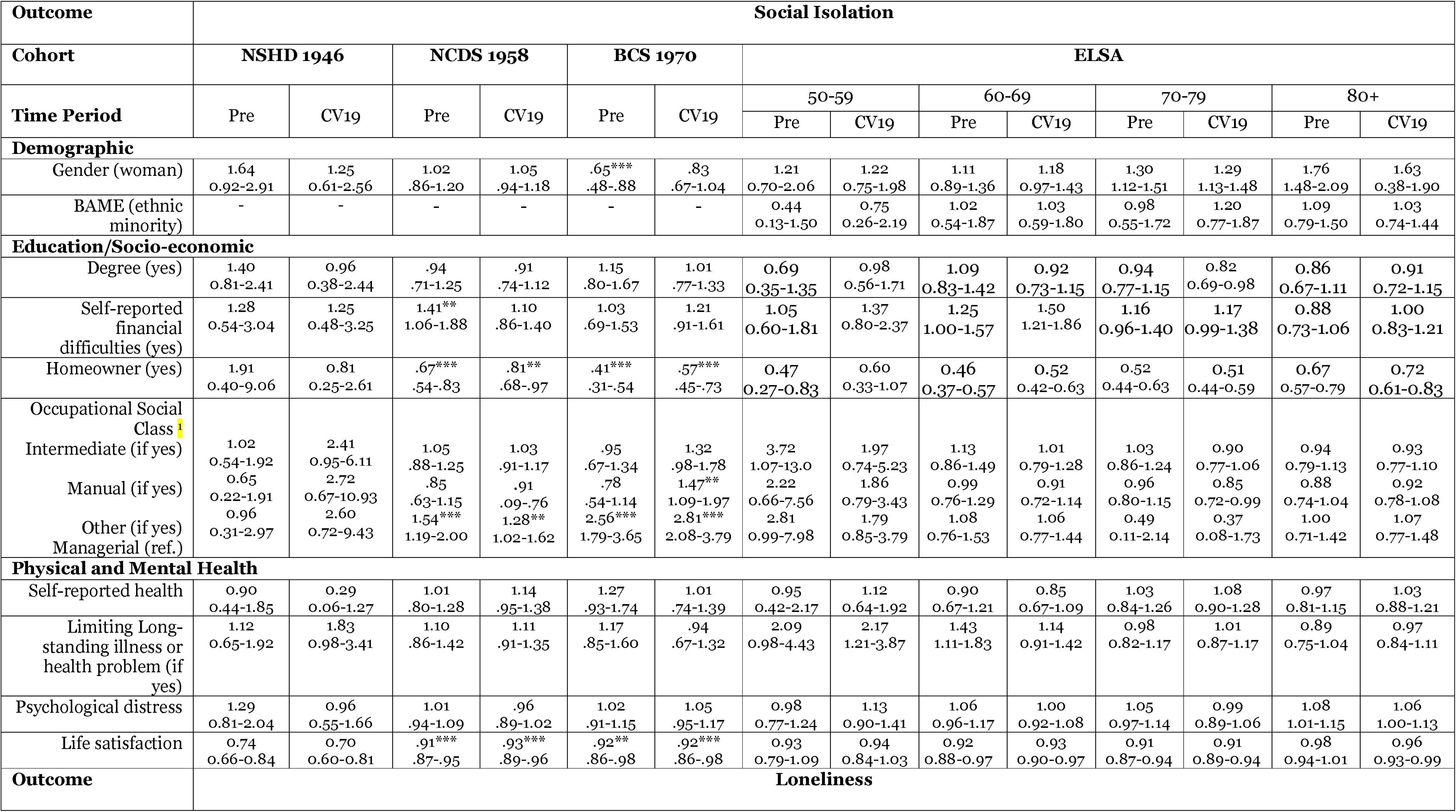
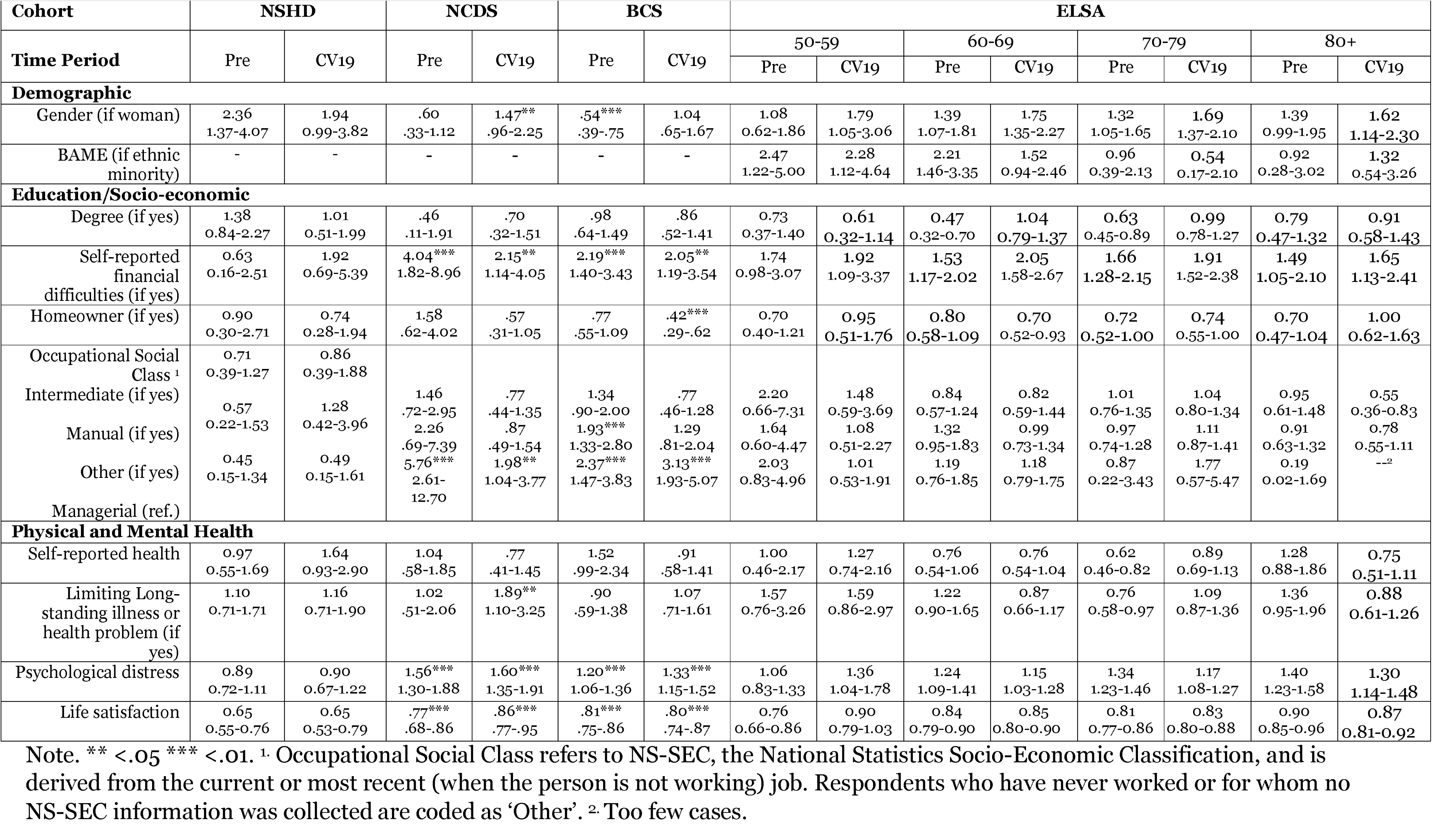
Results from modified Poisson regression models for social isolation and loneliness prior to and during the COVID-19 pandemic including relative risk (RR) and 95% Confidence Intervals (95% CI)

Correlates of greater social isolation included female gender, manual occupational social class, self-reported financial difficulties, not being a homeowner, longstanding illness and lower life satisfaction. There were no notable differences in the effect sizes of these associations in the pre-pandemic and lockdown periods.

Correlates of greater loneliness included female gender, not having degree level education, manual occupational social class, self-reported financial difficulties, not being a homeowner, longstanding illness and greater psychological distress and lower life satisfaction. There were no notable differences in the effect sizes of these associations in the pre-pandemic and lockdown periods.

## Discussion and Implications

This study provides a conceptual and empirical contribution, presenting the inter-relationship between social isolation and loneliness before and during the pandemic, with its unprecedented social restrictions. By using data from multiple successive birth cohorts, alongside several age-bands derived from a multi-generational ageing cohort, we were also able to examine cross-cohort differences in experiences during the early stages of COVID-19 and distinguish age or cohort effects from period effects related to the pandemic. Furthermore, we present the demographic, socio-economic, and health factors associated with experiences of social isolation and loneliness prior to and during the lockdown restrictions, adding to the literature that highlights the unequal burden of these experiences for females, those with greater economic precarity, including not owning a home, manual occupational social class, and greater financial stress.

We show support for these experiences as independent but related constructs (Hughes et al., 2004); however, our examination of demographic, socio-economic and health characteristics suggests mostly common correlates for these outcomes. Across all the datasets examined, and during both pre-pandemic and restricted periods of restrictions, more people reported being socially isolated than lonely based on our cut-off points. This maps onto findings from the European Social Survey collected before the pandemic, which indicated that 8.6 percent of the adult population had frequent feelings of loneliness whereas 20.8 percent were socially isolated (d’Hombres et al., 2021). When examining age-based differences, we observe higher levels of isolation at older ages. However, levels of loneliness were more stable across later life. When comparing different age groups, we see that the overlap between social isolation and loneliness is fairly consistent pre- and during the COVID-19 restrictions.

Examining the associations at the indicator-level, we find that the inter-relationships between indicators during the lockdown were less strongly connected compared to before the pandemic. This analysis suggests that there might have been an impact on how these concepts relate during restrictions, but these differences were not marked and are unlikely to indicate any fundamental differences in the conceptual links between loneliness and social isolation indicators during lockdown. The COVID-19 pandemic restrictions had specific effects on the prevalence of some of these indicators. For instance, remote contact increased and likely partially compensated for reductions in in-person contacts (Wels et al., 2023). There were also substantially fewer individuals in education and employment (Wels et al., 2022).

The study has several strengths, including the use of multiple data sources with slightly different designs and measures that permit the examination of consistency and replication of findings. However, there are limitations to note including the lack of availability of detailed measures of loneliness in the three birth cohorts before the pandemic, and data availability for relevant measures at certain timepoints (e.g., ELSA did not collect measures of face-to-face contact during the pandemic). We used data from participants who had responded before and during the pandemic surveys; this maintains a comparable sample, although those who dropped out might have been more isolated and in poorer health, potentially leading to some underestimation in the observed associations despite the use of sample and non-response weights in analyses.

By comparing data prior to and during enforced pandemic restrictions, we were able to provide insight into how these associations might vary in different contexts. In both these periods, we find similar associations between indicators of social isolation and loneliness, and with regard to other demographic, socioeconomic, and health correlates in both these periods. The number of study members reporting only isolation or loneliness, and the moderate overlap between older adults reporting both experiences indicate that these concepts are not transferable, and clarity in how they are conceptualised, operationalised and interpreted in quantitative research is necessary. This, in turn, will contribute to a better understanding of the role and consequences of social isolation and loneliness in older age, and inform how interventions might support different aspects of these outcomes in older adults.

The increases in social isolation observed in the pandemic highlight the need for efforts to encourage older people to (re)start hobbies, volunteer, and schedule time to meet up with friends and neighbours, as these activities can also lead to health and other psychological benefits (Fancourt et al., 2022). The findings that being female, economic precarity, and long-standing illness are stable correlates of isolation and loneliness indicate a need for structural changes and policies designed to reduce these inequalities in experiences of isolation and loneliness.

We would also like to highlight the implications of our findings for the currently widespread conflation of the two terms “loneliness” and “social isolation” in policy, and the over emphasis on loneliness within the UK context (e.g., campaign to end loneliness, loneliness ministers). Given many older adults experience high levels of social isolation, there should be greater emphasis on reducing social isolation through policy intervention rather than focusing on reducing individuals’ feelings of loneliness. Investigation of objective social isolation shifts the focus away from individuals and towards structural factors contributing greater isolation and the inequities in these experiences and contributing factors (Umberson et al., 2022). This can help to identify areas that are modifiable through targeted policy and intervention.

## Data Availability

All data produced are available upon request online at https://www.data-archive.ac.uk

## Acknowledgements

We are grateful to all the cohort members who took part in the NSHD, NCDS, BCS and ELSA cohorts analysed in this paper. We are also grateful to the survey and data management teams of these cohorts.

## Supplementary material

Supplementary file S1. Demographic, socio-economic and health characteristics of the analytic samples

Supplementary file S2. Variables harmonization

Supplementary file S3. Networks by age-bands in ELSA

