## Supplementary file S1 for "Examining the inter-relationships between social isolation and loneliness and their correlates among older British adults before and during the COVID-19 lockdown: evidence from four British longitudinal studies"

### **Supplementary file S1. Demographic, socio-economic and health characteristics of the analytic samples**

| **Variable** | **NSHD** | | **NCDS** | | **BCS** | | **ELSA** | |
| --- | --- | --- | --- | --- | --- | --- | --- | --- |
|  | **t-2** | **t-1** | **t-2** | **t-1** | **t-2** | **t-1** | **t-2** | **t-1** |
| **Sample Size** | **763** | | **3,851** | | **3,075** | | **4,728** | |
| **Demographic** | | | | | | | | |
| % Women | 48.3 (368) | | 51.8 (2,079) | | 49.6 (1,834) | | 52.7 (2,502) | |
| Mean Age (SD) | - | - | - | - | - | - | 66.5 (9.22) | 68.6 (9.15) |
| % BAME | - | - | - | - | - | - | 4.8 (115) | |
| **Education** | | | | | | | | |
| % Degree | 10.8 (82) | 10.8 (82) | 4.8 (224) | 4.8 (244) | 7.1 (327) | 7.2 (335) | 18.1 (923) | 21.7 (1,116) |
| **Socio-economic** | | | | | | | | |
| % Self-reported financial difficulties | 5.3 (40) | 2.8 (21) | 6.7 (193) | 6.7 (197) | 8.7 (205) | 6.0 (123) | 23.0 (774) | 18.8 (640) |
| % Homeowner (with/out mortgage) | 91.1 (695) | 96.5 (736) | 85.7 (3,447) | 86.7 (3,507) | 74.6 (2,546) | 74.6 (2,546) | 84.5 (3,956) | 84.5 (3,964) |
| Occupational status/social class  % Managerial  % Intermediate  % Manual  % Other | 10.0 (76)  69.3 (528)  11.7 (90)  9.0 (69) | 10.0 (76)  69.3 (528)  11.7 (90)  9.0 (69) | 42.6 (1,879)  33.5 (1,215)  10.8 (351)  13.2 (401) | 47.3 (2,094)  29.8 (1,075)  15.1 (478)  7.9 (191) | 45.2 (1,581)  18.6 (595)  20.7 (564)  15.4 (335) | 43.9 (1,567)  20.4 (620)  20.4 (535)  15.3 (353) | 29.3 (1,622)  22.4 (1,145)  34.1 (1,394)  14.2 (279) | 29.8 (1,623)  22.8 (1,158)  34.7 (1,411)  12.7 (248) |
| **Physical health** | | | | | | | | |
| Self-reported health -  % Poor/Fair | 6.8 (52) | 12.1 (92) | 14.5 (440) | 15.0 (432) | 16.0 (323) | 21.4 (434) | 24.3 (938) | 25.9 (999) |
| % Limiting long-standing illness/health problem | 17.1 (130) | 20.2 (154) | 12.1 (362) | 14.2 (429) | 18.2 (413) | 21.1 (498) | 31.8 (1,358) | 33.7 (1,465) |
| **Mental health** | | | | | | | | |
| Life Satisfaction –  Mean (SD) | 5.6 (1.33) | 5.6 (1.33) | 7.38 (1.66) | 7.37 (1.62) | 1.21 ( | 7.38 (1.67) | 7.28 (2.14) | 7.44 (2.24) |
| Psychological distress –  Mean (SD) | 1.21 (0.60) | 1.14 (0.42) | 1.45 (1.62) | 1.36 (1.74) | 1.85 (1.85) | 1.85 (1.85) | 1.31 (1.82) | 1.35 (1.81) |

Note. *Weighted %. Except for sex and ethnicity, all measures in each column refer to t-2 (time point before the most recent sweep before the COVID-19 survey) and t-1 (the most recent time point before the COVID-19 survey), where t represents the COVID-19 time point.*
