## Supplementary file S2 for "Examining the inter-relationships between social isolation and loneliness and their correlates among older British adults before and during the COVID-19 lockdown: evidence from four British longitudinal studies"

### **Supplementary file S2. Variables harmonization**

| ***Variables*** | ***Study*** | ***Variable code*** | ***Recoding if needed*** | ***Notes*** |
| --- | --- | --- | --- | --- |
| **Social Isolation** |  |  |  |  |
| ***Household Size* - total in the household including CM** |  |  |  |  |
|  | **NSHD NCDS BCS** | CW1_HHNUM | e.g., NCDS 9/11=8 | Recode to cap highest number in the household to ensure that anonymity is protects i.e., those with Ns under 10 |
|  | **ELSA** | CvNumP (at Covid Wave1) - How many people (including you) are currently living in the residence you are staying? |  | Values in sample range from 0 to 9 |
| ***Living Alone* 0=Not Living Alone;1=Living Alone** |  |  |  |  |
|  | **NSHD NCDS BCS** | CW1_HHNUM | e.g., NCDS 1/8=1 |  |
|  | **ELSA** | CvNumP | 2/max=0 |  |
| ***Family Network* - Children, in household, outside of household, any. 0=No Children; 1=Any Children** |  |  |  |  |
|  | **NSHD NCDS BCS** | CW1_HHNUMWH_2 CW1_ANYCHNL | 1=1; 2=0, gen (HH_CHILD_CV19) 1=1; 2=0, gen (FR_CHILD_CV19) gen ANY_CHILD_CV19 = . replace ANY_CHILD_CV19 = 0 if HH_CHILD_CV19==0 & FR_CHILD_CV19==0 replace ANY_CHILD_CV19 = 1 if HH_CHILD_CV19==1 \| FR_CHILD_CV19==1 | Generate whatever is available if only within household, fine, but use the same naming convention so it is clear |
|  | **ELSA** | Not asked in Covid Waves - we use pre-pandemic data |  |  |
| ***Family Network* - Partner, in the household, outside of household, any. 0=No Partner; 1=Any Partner** |  |  |  |  |
|  | **NSHD NCDS BCS** | CW1_HHNUMWH_1 CW1_OTHRELA | 1=1; 2=0, gen (HH_CHILD_CV19) 1=1; 2=0, gen (FR_CHILD_CV19) gen ANY_CHILD_CV19 = . replace ANY_CHILD_CV19 = 0 if HH_CHILD_CV19==0 & FR_CHILD_CV19==0 replace ANY_CHILD_CV19 = 1 if HH_CHILD_CV19==1 \| FR_CHILD_CV19==1 | Generate whatever is available if only within household, fine, but use the same naming convention so it is clear |
|  | **ELSA** | PartInHH (derived) - Relationship with other members in family is 'Husband/ Wife/ Partner'. 1=Yes, -1=Not applicable (only 1 person in HH) | 0 if PartInHH=-1 |  |
| ***Family and Friends* - weekly in person contact with friends/relatives. 0=No Weekly Contact; 1=Weekly Contact** |  |  |  |  |
|  | **NSHD NCDS BCS** | CW1_SCON1 CW1_SCON2 CW1_SCON3 | 1/4=1; 5=0, gen (FR_FF_FRQ_CV19) recode CW1_SCON2 CW1_SCON3 (-8=.) (1/4=1) (5=0), gen (FR_REM_FRQ_CV19_1 FR_REM_FRQ_CV19_2) gen FR_REM_FRQ_CV19 = . replace FR_REM_FRQ_CV19 = 0 if FR_REM_FRQ_CV19_1==0 & FR_REM_FRQ_CV19_2==0 replace FR_REM_FRQ_CV19 = 1 if FR_REM_FRQ_CV19_1==1 \| FR_REM_FRQ_CV19_2==1 gen ANY_FR_FRQ_CV19 = . replace ANY_FR_FRQ_CV19 = 0 if FR_FF_FRQ_CV19==0 & FR_REM_FRQ_CV19==0 replace ANY_FR_FRQ_CV19 = 1 if FR_FF_FRQ_CV19==1 \| FR_REM_FRQ_CV19==1 | Generate whatever is available if only within household, fine, but use the same naming convention so it is clear |
|  | **ELSA** | N/A |  |  |
| ***Family and Friends* - weekly remote contact with friends/relatives. 0=No Weekly Contact; 1=Weekly Contact** |  |  |  |  |
|  | **NSHD NCDS BCS** | CW1_SCON1 CW1_SCON2 CW1_SCON3 | 1/4=1; 5=0, gen (FR_FF_FRQ_CV19) recode CW1_SCON2 CW1_SCON3 (-8=.) (1/4=1) (5=0), gen (FR_REM_FRQ_CV19_1 FR_REM_FRQ_CV19_2) gen FR_REM_FRQ_CV19 = . replace FR_REM_FRQ_CV19 = 0 if FR_REM_FRQ_CV19_1==0 & FR_REM_FRQ_CV19_2==0 replace FR_REM_FRQ_CV19 = 1 if FR_REM_FRQ_CV19_1==1 \| FR_REM_FRQ_CV19_2==1 gen ANY_FR_FRQ_CV19 = . replace ANY_FR_FRQ_CV19 = 0 if FR_FF_FRQ_CV19==0 & FR_REM_FRQ_CV19==0 replace ANY_FR_FRQ_CV19 = 1 if FR_FF_FRQ_CV19==1 \| FR_REM_FRQ_CV19==1 | Generate whatever is available if only within household, fine, but use the same naming convention so it is clear |
|  | **ELSA** | FAM: In the past month, how often have you done the following with any of your immediate family (parents, children, grandchildren and brothers and sisters), not counting any who live with you? FRD: In the past month, how often have you done the following with other relatives and/or friends?   1. Speak on the phone -- 1.Daily; 2.Three to Six times a week; 3.Once/Twice a week; 4.Less than once a week or never  2. Write/ email -- 1.Daily; 2.Three to Six times a week; 3.Once/Twice a week; 4.Less than once a week or never  3. Send/ receive texts -- 1.Daily; 2.Three to Six times a week; 3.Once/Twice a week; 4.Less than once a week or never | 0 (no weekly contact) if (fam1==4 & fam2==4 & fam3==4) & (frd1==4& frd2==4 & frd3==4) |  |
| ***Education and employment* - neither in education nor employment. 0=Neither in Education Nor Employment; 1=In Education or Employment** |  |  |  |  |
|  | **NSHD NCDS BCS** | CW1_ECONACTIVITYD | 1=1; 2/4=0; 5=1; 6/9=0; 10=1; 11/13=0 |  |
|  | **ELSA** | CvPstd. Which of the following would you say best describes your current situation?  1.Retired; 2.Employed; 3.Paid/unpaid leave from employment (including furlough); 4.Self-employed and currently working; 5.Self-employed but not currently working; 6.Unemployed; 7.Permanently sick or disabled; 8.Looking after home or family | 1 if CvPstd==2 \| CvPstd_w1==4 0 for all other categories |  |
| ***Community engagement* - membership to club/organisation or volunteering. 0=No Community Engagement; 1=Community Engagement** |  |  |  |  |
|  | **NSHD NCDS BCS** |  | 1/4=1; 5=0 |  |
|  | **ELSA** | CvVolun. Have you changed the frequency you take part in voluntary work due to the coronavirus outbreak? 1.Yes, stopped completely; 2.Yes, less than before; 3.Yes, more than before; 4.No, about the same; 5.I did not volunteer previously | 1 if CvVolun==2 OR 3 OR 4 0 if CvVolun==1 OR 5 |  |
| ***Overall social isolation indicator* 0-6; 0=Not Socially Isolated; 1=Socially Isolated** |  |  |  |  |
|  | **NSHD NCDS BCS** |  | gen SI_CV19 = . replace SI_CV19 = 0 if HH_ALONE_CV19==1 & FR_FF_FRQ_CV19==0 & EE_SIZ_CV19_DUR==0 & /// COM_FRQ_CV19==0 & COM_VOL_CV19==0 replace SI_CV19 = 1 if HH_ALONE_CV19==0 \| FR_FF_FRQ_CV19==1 \| EE_SIZ_CV19_DUR==1 \| /// COM_FRQ_CV19==1 \| COM_VOL_CV19==1 |  |
| **Loneliness** |  |  |  |  |
| ***UCLA Loneliness Scale* - total score and binary 0=Not Lonely; 1=Lonely** |  |  |  |  |
|  | **NSHD NCDS BCS** |  | recode CW1_LONELY_1 CW1_LONELY_2 CW1_LONELY_3 CW1_LONELY_4 (-8=.), gen (LONE_CV19_1 LONE_CV19_2 LONE_CV19_3 LONE_CV19_4) egen LONE_TOTAL_CV19 = rowtotal(LONE_CV19_1 LONE_CV19_2 LONE_CV19_3) , missing gen LONE_CV19_01 = . replace LONE_CV19_01 = 0 if LONE_TOTAL_CV19==3 \| LONE_TOTAL_CV19==4 \| LONE_TOTAL_CV19==5 replace LONE_CV19_01 = 1 if LONE_TOTAL_CV19==6 \| LONE_TOTAL_CV19==7 \| LONE_TOTAL_CV19==8 \| LONE_TOTAL_CV19==9 gen LONE_CV19_4_01 = . replace LONE_CV19_4_01 = 0 if LONE_CV19_4==1 replace LONE_CV19_4_01 = 1 if LONE_CV19_4==2 \| LONE_CV19_4==3 |  |
|  | **ELSA** | A. How often do you feel you lack companionship? B. How often do you feel left out? C. How often do you feel isolated from others? D. How often do you feel lonely?   --- 1.Harldly Ever or Never; 2.Some of the time; 3.Often | For each question, 'lonely' if ==3  For UCLA total score: (mean of all scores)*3 If value is equal or greater than 6 --> Lonely |  |
