## Supplementary figures and images for "Examining the inter-relationships between social isolation and loneliness and their correlates among older British adults before and during the COVID-19 lockdown: evidence from four British longitudinal studies"

### Supplementary file S3

#### **Supplementary file S3. Networks by age-bands in ELSA**

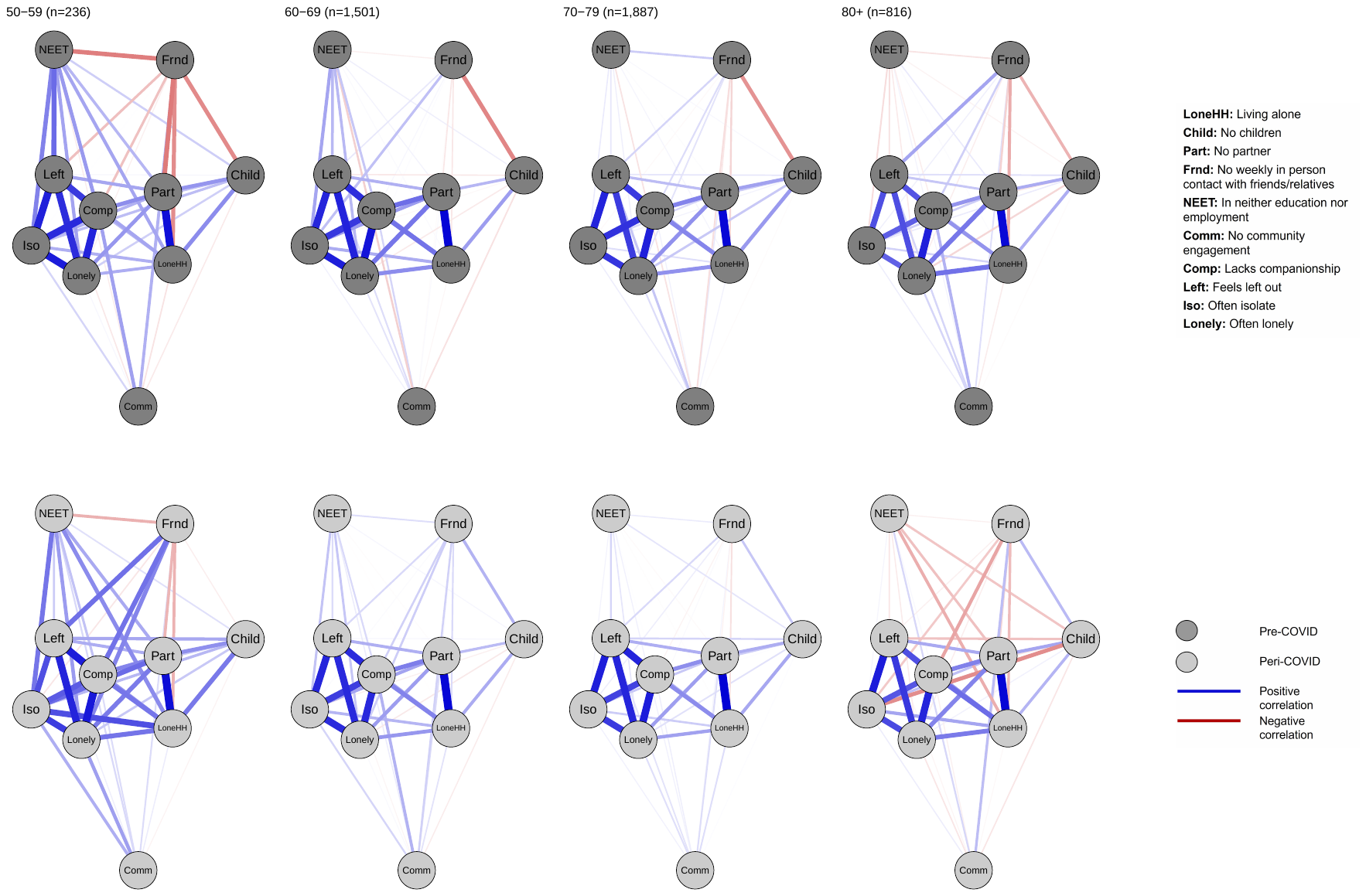
