## Supplementary file S4 for "Examining the inter-relationships between social isolation and loneliness and their correlates among older British adults before and during the COVID-19 lockdown: evidence from four British longitudinal studies"

### **Supplementary file S4. Tetrachoric correlation matrices**

#### NCDS pre-COVID

|  | LoneHH | Child | Part | Frnd | NEET | Comm | Lonely |
| --- | --- | --- | --- | --- | --- | --- | --- |
| LoneHH | 1 | 0.589983 | 0.981843 | -0.02602 | 0.042372 | 0.076241 | 0.135251 |
| Child | 0.589983 | 1 | 0.457515 | 0.073806 | -0.03365 | 0.025242 | 0.056223 |
| Part | 0.981843 | 0.457515 | 1 | -0.00374 | 0.098771 | 0.037865 | 0.091211 |
| Frnd | -0.02602 | 0.073806 | -0.00374 | 1 | -0.14037 | 0.12262 | 0.115433 |
| NEET | 0.042372 | -0.03365 | 0.098771 | -0.14037 | 1 | 0.039855 | 0.411054 |
| Comm | 0.076241 | 0.025242 | 0.037865 | 0.12262 | 0.039855 | 1 | 0.060861 |
| Lonely | 0.135251 | 0.056223 | 0.091211 | 0.115433 | 0.411054 | 0.060861 | 1 |

#### NCDS COVID

|  | LoneHH | Child | Part | Frnd | NEET | Comm | Lonely |
| --- | --- | --- | --- | --- | --- | --- | --- |
| LoneHH | 1 | 0.437798 | 0.751395 | -0.03528 | -0.08684 | 0.060672 | 0.317671 |
| Child | 0.437798 | 1 | 0.378367 | 0.01977 | 0.097583 | 0.052744 | 0.062364 |
| Part | 0.751395 | 0.378367 | 1 | -0.07829 | -0.01609 | 0.082119 | 0.328169 |
| Frnd | -0.03528 | 0.01977 | -0.07829 | 1 | -0.05501 | 0.21568 | 0.016342 |
| NEET | -0.08684 | 0.097583 | -0.01609 | -0.05501 | 1 | 0.006817 | 0.196053 |
| Comm | 0.060672 | 0.052744 | 0.082119 | 0.21568 | 0.006817 | 1 | 0.124797 |
| Lonely | 0.317671 | 0.062364 | 0.328169 | 0.016342 | 0.196053 | 0.124797 | 1 |

#### BCS pre-COVID

|  | LoneHH | Child | Part | Frnd | NEET | Comm | Lonely |
| --- | --- | --- | --- | --- | --- | --- | --- |
| LoneHH | 1 | 0.397456 | 0.602432 | -0.05752 | 0.408745 | 0.106978 | 0.40521 |
| Child | 0.397456 | 1 | 0.185457 | 0.098693 | 0.068008 | 0.04436 | 0.016794 |
| Part | 0.602432 | 0.185457 | 1 | -0.13935 | 0.351058 | 0.080645 | 0.283281 |
| Frnd | -0.05752 | 0.098693 | -0.13935 | 1 | -0.01185 | 0.158659 | 0.007892 |
| NEET | 0.408745 | 0.068008 | 0.351058 | -0.01185 | 1 | 0.227435 | 0.3624 |
| Comm | 0.106978 | 0.04436 | 0.080645 | 0.158659 | 0.227435 | 1 | 0.286927 |
| Lonely | 0.40521 | 0.016794 | 0.283281 | 0.007892 | 0.3624 | 0.286927 | 1 |

#### BCS COVID

|  | LoneHH | Child | Part | Frnd | NEET | Comm | Lonely |
| --- | --- | --- | --- | --- | --- | --- | --- |
| LoneHH | 1 | 0.540456 | 0.607044 | -0.12761 | 0.144962 | 0.064402 | 0.449424 |
| Child | 0.540456 | 1 | 0.326816 | 0.053917 | 0.020345 | 0.093774 | 0.13131 |
| Part | 0.607044 | 0.326816 | 1 | -0.01306 | 0.183432 | 0.032101 | 0.405652 |
| Frnd | -0.12761 | 0.053917 | -0.01306 | 1 | 0.053566 | 0.161491 | 0.058226 |
| NEET | 0.144962 | 0.020345 | 0.183432 | 0.053566 | 1 | 0.117307 | 0.385055 |
| Comm | 0.064402 | 0.093774 | 0.032101 | 0.161491 | 0.117307 | 1 | 0.251157 |
| Lonely | 0.449424 | 0.13131 | 0.405652 | 0.058226 | 0.385055 | 0.251157 | 1 |

#### ELSA Age 50 Pre-COVID

|  | LoneHH | Child | Part | Frnd | NEET | Comm | Comp | Left | Iso | Lonely |
| --- | --- | --- | --- | --- | --- | --- | --- | --- | --- | --- |
| LoneHH | 1 | 0.451849 | 0.939875 | -0.45286 | 0.353296 | -0.10794 | 0.276963 | 0.354281 | 0.314474 | 0.320358 |
| Child | 0.451849 | 1 | 0.414935 | -0.45795 | 0.190906 | -0.08095 | 0.244248 | 0.036647 | 0.259694 | 0.187217 |
| Part | 0.939875 | 0.414935 | 1 | -0.51836 | 0.287206 | 0.045539 | 0.447002 | 0.350625 | 0.304802 | 0.40931 |
| Frnd | -0.45286 | -0.45795 | -0.51836 | 1 | -0.50598 | 0.31161 | -0.24449 | -0.24965 | 0.00585 | -0.06043 |
| NEET | 0.353296 | 0.190906 | 0.287206 | -0.50598 | 1 | -0.06887 | 0.400835 | 0.582055 | 0.489074 | 0.360708 |
| Comm | -0.10794 | -0.08095 | 0.045539 | 0.31161 | -0.06887 | 1 | 0.374102 | -0.09162 | 0.16569 | 0.032271 |
| Comp | 0.276963 | 0.244248 | 0.447002 | -0.24449 | 0.400835 | 0.374102 | 1 | 0.774116 | 0.83787 | 0.863883 |
| Left | 0.354281 | 0.036647 | 0.350625 | -0.24965 | 0.582055 | -0.09162 | 0.774116 | 1 | 0.874752 | 0.823307 |
| Iso | 0.314474 | 0.259694 | 0.304802 | 0.00585 | 0.489074 | 0.16569 | 0.83787 | 0.874752 | 1 | 0.890673 |
| Lonely | 0.320358 | 0.187217 | 0.40931 | -0.06043 | 0.360708 | 0.032271 | 0.863883 | 0.823307 | 0.890673 | 1 |

ELSA Age 60 Pre-COVID

|  | LoneHH | Child | Part | Frnd | NEET | Comm | Comp | Left | Iso | Lonely |
| --- | --- | --- | --- | --- | --- | --- | --- | --- | --- | --- |
| LoneHH | 1 | 0.383085 | 0.978913 | -0.09568 | 0.01911 | -0.03119 | 0.549765 | 0.235998 | 0.323824 | 0.452652 |
| Child | 0.383085 | 1 | 0.287047 | -0.47519 | 0.062498 | -0.11484 | -0.00582 | -0.08846 | -0.02812 | -0.08428 |
| Part | 0.978913 | 0.287047 | 1 | -0.09239 | 0.04595 | 0.024217 | 0.568407 | 0.308607 | 0.38639 | 0.535454 |
| Frnd | -0.09568 | -0.47519 | -0.09239 | 1 | -0.06384 | -0.01628 | 0.031976 | 0.091371 | 0.2473 | 0.011792 |
| NEET | 0.01911 | 0.062498 | 0.04595 | -0.06384 | 1 | -0.18456 | 0.222777 | 0.279074 | 0.351702 | 0.276801 |
| Comm | -0.03119 | -0.11484 | 0.024217 | -0.01628 | -0.18456 | 1 | 0.163964 | 0.135114 | 0.052945 | 0.110457 |
| Comp | 0.549765 | -0.00582 | 0.568407 | 0.031976 | 0.222777 | 0.163964 | 1 | 0.822769 | 0.781851 | 0.92259 |
| Left | 0.235998 | -0.08846 | 0.308607 | 0.091371 | 0.279074 | 0.135114 | 0.822769 | 1 | 0.854579 | 0.867267 |
| Iso | 0.323824 | -0.02812 | 0.38639 | 0.2473 | 0.351702 | 0.052945 | 0.781851 | 0.854579 | 1 | 0.883253 |
| Lonely | 0.452652 | -0.08428 | 0.535454 | 0.011792 | 0.276801 | 0.110457 | 0.92259 | 0.867267 | 0.883253 | 1 |

#### ELSA Age 70 pre-COVID

|  | LoneHH | Child | Part | Frnd | NEET | Comm | Comp | Left | Iso | Lonely |
| --- | --- | --- | --- | --- | --- | --- | --- | --- | --- | --- |
| LoneHH | 1 | 0.399645 | 0.974446 | -0.16624 | 0.04001 | 0.052877 | 0.47768 | 0.265956 | 0.128811 | 0.430595 |
| Child | 0.399645 | 1 | 0.369355 | -0.42353 | 0.02554 | -0.01641 | 0.209588 | 0.099042 | 0.045297 | 0.216326 |
| Part | 0.974446 | 0.369355 | 1 | -0.09259 | 0.012459 | 0.088588 | 0.485734 | 0.262194 | 0.1555 | 0.429922 |
| Frnd | -0.16624 | -0.42353 | -0.09259 | 1 | 0.198287 | -0.12825 | 0.136949 | 0.165375 | -0.02239 | 0.111558 |
| NEET | 0.04001 | 0.02554 | 0.012459 | 0.198287 | 1 | -0.10868 | -0.00156 | 0.095476 | 0.080671 | 0.061863 |
| Comm | 0.052877 | -0.01641 | 0.088588 | -0.12825 | -0.10868 | 1 | 0.053688 | 0.228344 | 0.068298 | 0.138214 |
| Comp | 0.47768 | 0.209588 | 0.485734 | 0.136949 | -0.00156 | 0.053688 | 1 | 0.772677 | 0.806247 | 0.879767 |
| Left | 0.265956 | 0.099042 | 0.262194 | 0.165375 | 0.095476 | 0.228344 | 0.772677 | 1 | 0.863761 | 0.766242 |
| Iso | 0.128811 | 0.045297 | 0.1555 | -0.02239 | 0.080671 | 0.068298 | 0.806247 | 0.863761 | 1 | 0.79257 |
| Lonely | 0.430595 | 0.216326 | 0.429922 | 0.111558 | 0.061863 | 0.138214 | 0.879767 | 0.766242 | 0.79257 | 1 |

ELSA Age 80 pre-COVID

|  | LoneHH | Child | Part | Frnd | NEET | Comm | Comp | Left | Iso | Lonely |
| --- | --- | --- | --- | --- | --- | --- | --- | --- | --- | --- |
| LoneHH | 1 | 0.320526 | 0.966707 | -0.3238 | 0.01714 | -0.07537 | 0.420509 | 0.164338 | 0.177097 | 0.558602 |
| Child | 0.320526 | 1 | 0.328673 | -0.2977 | -0.04835 | -0.01623 | -0.09973 | 0.121302 | 0.078793 | -0.15653 |
| Part | 0.966707 | 0.328673 | 1 | -0.17532 | 0.080824 | -0.00057 | 0.418046 | 0.223032 | 0.195403 | 0.534549 |
| Frnd | -0.3238 | -0.2977 | -0.17532 | 1 | -0.10871 | 0.12524 | 0.189284 | 0.353869 | 0.060397 | -0.06802 |
| NEET | 0.01714 | -0.04835 | 0.080824 | -0.10871 | 1 | 0.045108 | -0.069 | -0.11976 | -0.13262 | -0.08319 |
| Comm | -0.07537 | -0.01623 | -0.00057 | 0.12524 | 0.045108 | 1 | 0.300684 | 0.164827 | 0.065193 | 0.166479 |
| Comp | 0.420509 | -0.09973 | 0.418046 | 0.189284 | -0.069 | 0.300684 | 1 | 0.671157 | 0.630271 | 0.873776 |
| Left | 0.164338 | 0.121302 | 0.223032 | 0.353869 | -0.11976 | 0.164827 | 0.671157 | 1 | 0.692109 | 0.692551 |
| Iso | 0.177097 | 0.078793 | 0.195403 | 0.060397 | -0.13262 | 0.065193 | 0.630271 | 0.692109 | 1 | 0.619083 |
| Lonely | 0.558602 | -0.15653 | 0.534549 | -0.06802 | -0.08319 | 0.166479 | 0.873776 | 0.692551 | 0.619083 | 1 |

#### ELSA Age 50 COVID

|  | LoneHH | Child | Part | Frnd | NEET | Comm | Comp | Left | Iso | Lonely |
| --- | --- | --- | --- | --- | --- | --- | --- | --- | --- | --- |
| LoneHH | 1 | 0.526352 | 0.945342 | -0.30334 | 0.453324 | 0.075848 | 0.535604 | 0.430429 | 0.690622 | 0.546878 |
| Child | 0.526352 | 1 | 0.434588 | -0.06617 | 0.11973 | -0.04773 | 0.206936 | 0.346076 | 0.282863 | 0.215577 |
| Part | 0.945342 | 0.434588 | 1 | -0.29054 | 0.335664 | 0.014807 | 0.483178 | 0.321416 | 0.579576 | 0.516607 |
| Frnd | -0.30334 | -0.06617 | -0.29054 | 1 | -0.30605 | 0.137356 | 0.551555 | 0.562314 | -0.09705 | 0.477383 |
| NEET | 0.453324 | 0.11973 | 0.335664 | -0.30605 | 1 | 0.150407 | 0.240969 | 0.202022 | 0.53946 | 0.392409 |
| Comm | 0.075848 | -0.04773 | 0.014807 | 0.137356 | 0.150407 | 1 | 0.138004 | 0.064083 | 0.318021 | 0.374614 |
| Comp | 0.535604 | 0.206936 | 0.483178 | 0.551555 | 0.240969 | 0.138004 | 1 | 0.890407 | 0.641802 | 0.907804 |
| Left | 0.430429 | 0.346076 | 0.321416 | 0.562314 | 0.202022 | 0.064083 | 0.890407 | 1 | 0.690864 | 0.85969 |
| Iso | 0.690622 | 0.282863 | 0.579576 | -0.09705 | 0.53946 | 0.318021 | 0.641802 | 0.690864 | 1 | 0.760406 |
| Lonely | 0.546878 | 0.215577 | 0.516607 | 0.477383 | 0.392409 | 0.374614 | 0.907804 | 0.85969 | 0.760406 | 1 |

ELSA Age 60 COVID

|  | LoneHH | Child | Part | Frnd | NEET | Comm | Comp | Left | Iso | Lonely |
| --- | --- | --- | --- | --- | --- | --- | --- | --- | --- | --- |
| LoneHH | 1 | 0.330398 | 0.973276 | 0.144898 | -0.00581 | 0.109869 | 0.487175 | 0.319536 | 0.289733 | 0.407375 |
| Child | 0.330398 | 1 | 0.266736 | 0.29848 | 0.03422 | -0.07286 | -0.01317 | 0.035607 | -0.06055 | -0.08771 |
| Part | 0.973276 | 0.266736 | 1 | 0.031978 | -0.01171 | 0.141829 | 0.507257 | 0.313698 | 0.355906 | 0.382725 |
| Frnd | 0.144898 | 0.29848 | 0.031978 | 1 | 0.127674 | 0.179484 | 0.183727 | 0.150305 | 0.016903 | 0.197825 |
| NEET | -0.00581 | 0.03422 | -0.01171 | 0.127674 | 1 | 0.074739 | 0.149956 | 0.138083 | 0.235699 | 0.223644 |
| Comm | 0.109869 | -0.07286 | 0.141829 | 0.179484 | 0.074739 | 1 | 0.302476 | -0.03924 | 0.162769 | 0.024973 |
| Comp | 0.487175 | -0.01317 | 0.507257 | 0.183727 | 0.149956 | 0.302476 | 1 | 0.834481 | 0.761324 | 0.842114 |
| Left | 0.319536 | 0.035607 | 0.313698 | 0.150305 | 0.138083 | -0.03924 | 0.834481 | 1 | 0.853745 | 0.838118 |
| Iso | 0.289733 | -0.06055 | 0.355906 | 0.016903 | 0.235699 | 0.162769 | 0.761324 | 0.853745 | 1 | 0.834776 |
| Lonely | 0.407375 | -0.08771 | 0.382725 | 0.197825 | 0.223644 | 0.024973 | 0.842114 | 0.838118 | 0.834776 | 1 |

ELSA Age 70 COVID

|  | LoneHH | Child | Part | Frnd | NEET | Comm | Comp | Left | Iso | Lonely |
| --- | --- | --- | --- | --- | --- | --- | --- | --- | --- | --- |
| LoneHH | 1 | 0.345508 | 0.994649 | -0.12214 | -0.00807 | 0.129009 | 0.450973 | 0.29103 | 0.199965 | 0.369476 |
| Child | 0.345508 | 1 | 0.321967 | 0.220415 | 0.102659 | 0.010607 | 0.19631 | 0.169037 | 0.008588 | 0.118528 |
| Part | 0.994649 | 0.321967 | 1 | -0.05076 | -0.0209 | 0.134498 | 0.443447 | 0.318151 | 0.217263 | 0.380893 |
| Frnd | -0.12214 | 0.220415 | -0.05076 | 1 | 0.078377 | 0.043453 | -0.04835 | 0.149143 | 0.0684 | 0.019326 |
| NEET | -0.00807 | 0.102659 | -0.0209 | 0.078377 | 1 | 0.056355 | 0.004408 | 0.162927 | 0.047568 | 0.096965 |
| Comm | 0.129009 | 0.010607 | 0.134498 | 0.043453 | 0.056355 | 1 | 0.067263 | 0.108393 | 0.053713 | 0.127339 |
| Comp | 0.450973 | 0.19631 | 0.443447 | -0.04835 | 0.004408 | 0.067263 | 1 | 0.857882 | 0.742761 | 0.886924 |
| Left | 0.29103 | 0.169037 | 0.318151 | 0.149143 | 0.162927 | 0.108393 | 0.857882 | 1 | 0.880747 | 0.854633 |
| Iso | 0.199965 | 0.008588 | 0.217263 | 0.0684 | 0.047568 | 0.053713 | 0.742761 | 0.880747 | 1 | 0.770768 |
| Lonely | 0.369476 | 0.118528 | 0.380893 | 0.019326 | 0.096965 | 0.127339 | 0.886924 | 0.854633 | 0.770768 | 1 |

#### ELSA Age 80 COVID

|  | LoneHH | Child | Part | Frnd | NEET | Comm | Comp | Left | Iso | Lonely |
| --- | --- | --- | --- | --- | --- | --- | --- | --- | --- | --- |
| LoneHH | 1 | 0.292347 | 0.978011 | -0.2649 | -0.31828 | -0.02852 | 0.576592 | 0.424378 | 0.3541 | 0.522757 |
| Child | 0.292347 | 1 | 0.351486 | 0.269162 | -0.22576 | -0.09649 | -0.10341 | -0.23734 | -0.44715 | 0.054253 |
| Part | 0.978011 | 0.351486 | 1 | -0.15908 | -0.24799 | -0.06333 | 0.493014 | 0.337039 | 0.248731 | 0.48098 |
| Frnd | -0.2649 | 0.269162 | -0.15908 | 1 | -0.05845 | 0.25755 | -0.35474 | -0.0579 | -0.20985 | -0.06607 |
| NEET | -0.31828 | -0.22576 | -0.24799 | -0.05845 | 1 | 0.155005 | 0.026979 | -0.10043 | 0.040525 | -0.07779 |
| Comm | -0.02852 | -0.09649 | -0.06333 | 0.25755 | 0.155005 | 1 | -0.05811 | -0.03848 | 0.070941 | -0.09663 |
| Comp | 0.576592 | -0.10341 | 0.493014 | -0.35474 | 0.026979 | -0.05811 | 1 | 0.689218 | 0.663719 | 0.857553 |
| Left | 0.424378 | -0.23734 | 0.337039 | -0.0579 | -0.10043 | -0.03848 | 0.689218 | 1 | 0.889344 | 0.781213 |
| Iso | 0.3541 | -0.44715 | 0.248731 | -0.20985 | 0.040525 | 0.070941 | 0.663719 | 0.889344 | 1 | 0.709647 |
| Lonely | 0.522757 | 0.054253 | 0.48098 | -0.06607 | -0.07779 | -0.09663 | 0.857553 | 0.781213 | 0.709647 | 1 |
